# Air pollution control efficacy and health impacts: A global observed study from 2000 to 2016

**DOI:** 10.1101/2020.05.31.20118752

**Authors:** Chunlei Han, Rongbin Xu, Yajuan Zhang, Wenhua Yu, Shanshan Li, Zhongwen Zhang, Lidia Morawska, Jane Heyworth, Bin Jalaludin, Geoffrey Morgan, Guy Marks, Michael Abramson, Liwei Sun, Yuming Guo

## Abstract

**Background:** PM_2.5_ concentrations vary between countries with similar CO_2_ emissions, possibly due to differences in air pollution control efficacy. However, no indicator of the level of air pollution control efficacy has yet been developed. We aimed to develop such an indicator, and to evaluate its global and temporal distribution and its association with country-level health metrics.

**Method:** A novel indicator, ground level population-weighted average PM_2.5_ concentration per unit CO_2_ emission per capita (*PM*_2.5_/*CO*_2_, written as PC in abbreviation), was developed to assess country-specific air pollution control efficacy. We estimated and mapped the global average distribution of PC and PC changes during 2000–2016 across 196 countries. Pearson correlation coefficients and Generalized Additive Mixed Model (GAMM) were used to evaluate the relationship between PC and health metrics.

**Results:** PC varied by country with an inverse association with the economic development. PC showed an almost stable trend globally from 2000 to 2016 with the low income groups increased. The Pearson correlation coefficients between PC and life expectancy at birth (LE), Infant-mortality rate (IMR), Under-five mortality rate (U5MR) and logarithm of GDP per capita (LPGDP) were –0.566, 0.646, 0.659, –0.585 respectively (all P-values <0.001). Compared with PM_2.5_ or CO_2_, PC could explain more variation of LE, IMR and U5MR. The association between PC and health metrics was independent of GDP per capita.

**Conclusions:** PC might be a good indicator for air pollution control efficacy and was related to important health indicators. Our findings provide a new way to interpret health inequity across the globe from the point of air pollution control efficacy.

## 1. Introduction

Ambient air pollution is a major public health concern. Among all ambient air pollutants, the particulate matter with aerodynamic diameter ≤2.5um (PM_2.5_) is the most important one that poses significant adverse health impacts in both short-term and long-term[1,2]. At the same time, carbon dioxide (CO_2_) emissions have increased rapidly along with the rapid growth of economic development requiring more energy for transportation and energy consumption. As both ground level PM_2.5_ and CO_2_ are mainly caused by fossil-fuel combustion [3], there might be a relationship between CO_2_ emission and ground level PM_2.5_ concentration [4]. Studies conclude that actions to reduce greenhouse gas emissions often lead to co-benefits for air quality [5]. But interestingly, ground level PM_2.5_ concentrations are quite different across countries with similar CO_2_ emissions [4]. Many low- and middle-income countries (LMICs) face the dual pressure of reducing both ground level PM_2.5_ concentrations and CO_2_ emissions[5], while high income countries (HICs) have much lower ground level PM_2.5_ concentrations despite their high greenhouse gas emissions[6]. In other words, economically developed countries generally have lower ground level PM_2.5_ concentrations and relatively good air quality compared with economically developing countries, despite their similar or even higher CO_2_ emissions[7]. This fact suggests that different countries have different abilities to control ambient air pollution, even with similar CO_2_ emissions. An indicator to reflect the air pollution control efficacy may provide important information for policymakers, in order to achieve climate and air quality co-benefits and help guide environmental policy development and implementation [8].

The combustion sources of ground level PM_2.5_ concentrations are different across countries. Ground level PM_2.5_ concentrations are substantially from residential energy use such as heating and cooking in China, India, Bangladesh, Indonesia, Vietnam and Nepal; from traffic in Germany, the UK and the USA; from power generation in the USA, Russia, Korea, Turkey and the Middle East; from agriculture in Europe, Russia, Turkey, Korea, Japan and the Eastern USA[9]. Energy structure and environmental technology are both determinants of air pollution control efficacy. Environmental technological progress can enhance energy efficiency, thereby leading to reductions in ground level PM_2.5_ concentrations [4,10]. Developed countries may have more economic foundation to promote and apply technological innovation to reduce both CO_2_ emission and ground level PM_2.5_ concentration compared with developing countries. In developed countries such as North America and Europe, technological improvements in scrubbers on power plants, catalytic converters on motor vehicles, and increased development of non-fossil fuel based energy sources have reduced ground level PM_2.5_ concentrations [11]. Although emission reduction technologies play a role in improving air quality in economically developing countries like China [12], not all effective strategies are adopted due to the high cost[13].

Cleaner air due to air pollution reduction will improve human health[13]. Correspondingly, inequality in air pollution control efficacy contributes to human health inequality between countries[14]. An indicator of air pollution control efficacy could help identify the ground level air pollution concentration co-benefits of reducing emissions of CO_2_ [15]. The quantitative relationship between the air pollution control efficacy indicator and human health might provide important guidance for policymakers to reduce the disease burden due to ambient air pollution globally [4].

Currently, there is no indicator to reflect country level air pollution control efficacy. To fill the research gap, we aim to evaluate a potential novel indicator of air pollution control efficacy, by quantifying its global distribution and long-term trend, and by examining its relationship with health indicators. Monitoring such an indicator may assist policy makers to better manage climate change and air pollution problems simultaneously [5].

## 2. Materials and methods

### 2.1 Indicator

To capture air pollution control efficacy with CO_2_ emission, we proposed a novel indicator, ground level population weighted PM_2.5_ concentration per unit CO_2_ emission per capita (PC). A lower PC value generally indicates a higher air pollution control efficiency, meaning lower concentration of ground level PM_2.5_ with per unit of CO_2_ emission. The unit of PC is µg/m^3^ per tonne. PC is calculated as follows:

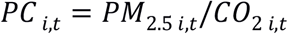

Here, i means the i^th^country or region, t means the t^th^ year.

### 2.2. Data collection

The spatial and temporal domain of our study included 196 countries from 2000 and 2016. Some regions like Greenland, Antarctica and some countries in Middle Africa were not included in the spatial map because of the missing data.

To develop the novel indicator of air pollution control efficacy, population-weighted ground level PM_2.5_ (PM_2.5_, µg/m^3^) and annual emissions of carbon dioxide per capita (CO_2_, tonne) for individual countries based on territorial CO_2_ emissions were sourced from the atmospheric composition analysis group, Global Carbon Project, Carbon Dioxide Information Analysis Centre (CDIAC), Gapminder and UN population estimates(see supplement for more details). PM_2.5_ in each country was represented by the population density weighted average value of all grids within the boundary of the country[16]. We transformed the original spatial resolution of this population density dataset into 0.1^°^ × 0.1^°^resolution according to the method described by Brauer et al[17].

To evaluate the association between PC and health, we collected data on several health outcomes. The first one is life expectancy at birth (LE, years), defined as the average number of years that a newborn could expect to live if he or she were to pass through life subject to the age-specific mortality rates of a given period. Children are more affected by air pollution and climate change [3,18]. It was reported that per 10 μg/m^3^ increases in PM_2.5_ concentration was related to 3.4% (95% CI: 1.7%–5.4%) infant and child under-five mortality[19]. Therefore,we included the health outcomes of infant-mortality rate (IMR, ‰) and under-five mortality rate (U5MR, ‰),which mean the number of infants dying before reaching one year of age and the number of babies that died before reaching age five per 1,000 live births in a given year. We obtained data of LE, IMR, U5MR from various sources including the United Nations (UN) Population Division, World Bank(WB), UN Inter-agency Group for Child Mortality Estimation, World Health Organization (WHO) (see supplement for more details).

Temperature and humidity are related to health [20] and country-level annual average Temperature at 2 meters (T2M,°C) and Specific Humidity at 2 Meters (QV2M, g water/kg dry air, g kg-1) were obtained from the National Aeronautics and Space Administration (NASA) (see supplement in details). GDP per capita (PGDP, U.S.$) in constant 2010 U.S. dollars came from WB and the Organization for Economic Co-operation and Development (OECD) (see supplement for more details).

### 2.3 Statistical Methods

Correlations between each two independent variables were examined by Pearson correlation coefficient. The Generalized Additive Mixed Model (GAMM) with a penalized spline smoothing function, a random intercept of country and spatial covariance structure, and a Gaussian link function, was used to evaluate the potential non-linear relationship between PC and health outcomes [21,22].

To ensure the results’ robustness, we excluded 5% observations with extreme large and small PC and kept the remaining 95% data in the middle for analyses. The model performance was expressed as adjusted R^2^. The GAMM was as following:

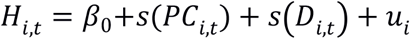

Here *H* represents the health outcome, which could be LE, IMR, or U5MR; i,t means the i^th^ country(i = 1 to 196) in the t^th^ (t = 2000 to 2016) year. β_0_ denotes the constant intercept; s(.) is the smoothing function realized by cubic spline with 4 degrees of freedom(df) in this study. *u_i_* is a random intercept for country i. D represents the covariates including PGDP, T2M, QV2M, PM_2.5_, CO_2_. The degrees of freedom (df) of the cubic spline function (CS) for each predictor was selected by minimizing the Akaike information criterion (AIC) of the model [23–25].

PC showed nonlinear correlation with health metrics as estimated in this paper, so here PC was modelled by a non-linear function. PGDP, T2M, QV2M were added to the models in the form of a natural cubic smooth function as their relationship with heath is often non-linear [26–28]. PM_2.5_ and CO_2_ were also included as covariates.

All statistical tests were two-sided, with a p-value of 0.05 as the indicator of the statistical significance. All analyses were performed using the R statistical software (version 3.2.2), including the R packages “ggplot2”, “dplyr”, “reldist” and “gamm4”.

## 3. Results

### 3.1 Descriptive results

The means of PM_2.5_ and CO_2_ were 21.52 (µg/m^3^) and 4.60 (tonne) respectively. PC was 74.24 (µg/m^3^ per tonne) on average with the considerable international variance from 0.14 (µg/m^3^ per tonne) in Australia (2010) to 2659.75 (µg/m^3^ per tonne) in Chad (2002). The average LE, IMR and U5MR were 68.94 years, 2.97 ‰ and 4.27‰, respectively. PGDP was 15541.76 (U.S.$) on average with a large range of 155795.00 (U.S.$). As for average temperature and humidity, T2M was18.33 (°C) and QV2M10.03 (g kg^−1^) (see Table 1). Generally, PC was lowest in high income groups, and then upper-middle income groups, lower-middle income groups, and highest in low income groups[29]. The mean, median, standard deviation and range of PC were increasing as the GDP per capita decreased (Table S1).

**Table 1.**
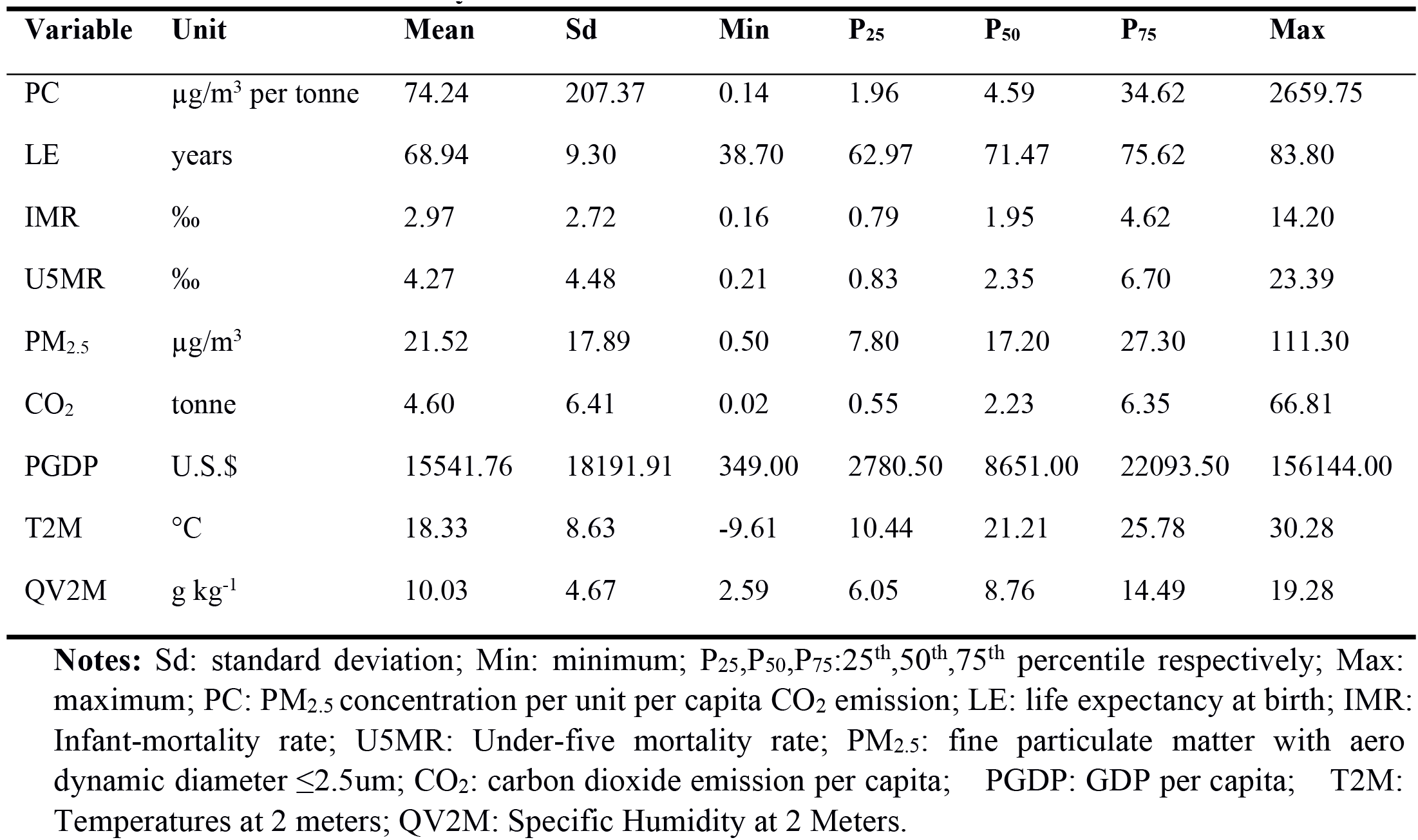
Summary statistics of all variables in 196 countries between 2000 and 2016.

| Variable | Unit | Mean | Sd | Min | P<sub>25</sub> | P<sub>50</sub> | P<sub>75</sub> | Max |
| --- | --- | --- | --- | --- | --- | --- | --- | --- |
| PC | µg/m<sup>3</sup> per tonne | 74.24 | 207.37 | 0.14 | 1.96 | 4.59 | 34.62 | 2659.75 |
| LE | years | 68.94 | 9.30 | 38.70 | 62.97 | 71.47 | 75.62 | 83.80 |
| IMR | ‰ | 2.97 | 2.72 | 0.16 | 0.79 | 1.95 | 4.62 | 14.20 |
| U5MR | ‰ | 4.27 | 4.48 | 0.21 | 0.83 | 2.35 | 6.70 | 23.39 |
| PM<sub>2.5</sub> | µg/m<sup>3</sup> | 21.52 | 17.89 | 0.50 | 7.80 | 17.20 | 27.30 | 111.30 |
| CO<sub>2</sub> | tonne | 4.60 | 6.41 | 0.02 | 0.55 | 2.23 | 6.35 | 66.81 |
| PGDP | U.S.\$ | 15541.76 | 18191.91 | 349.00 | 2780.50 | 8651.00 | 22093.50 | 156144.00 |
| T2M | °C | 18.33 | 8.63 | -9.61 | 10.44 | 21.21 | 25.78 | 30.28 |
| QV2M | g kg<sup>-1</sup> | 10.03 | 4.67 | 2.59 | 6.05 | 8.76 | 14.49 | 19.28 |

**Notes:** Sd: standard deviation; Min: minimum; P<sub>25</sub>, P<sub>50</sub>, P<sub>75</sub>: 25<sup>th</sup>, 50<sup>th</sup>, 75<sup>th</sup> percentile respectively; Max: maximum; PC: PM<sub>2.5</sub> concentration per unit per capita CO<sub>2</sub> emission; LE: life expectancy at birth; IMR: Infant-mortality rate; U5MR: Under-five mortality rate; PM<sub>2.5</sub>: fine particulate matter with aerodynamic diameter ≤2.5µm; CO<sub>2</sub>: carbon dioxide emission per capita; PGDP: GDP per capita; T2M: Temperatures at 2 meters; QV2M: Specific Humidity at 2 Meters.

### 3.2 Spatial and temporal variation of PC

The PC, PM_2.5_ and CO_2_ trends of the whole world, different income groups (high, upper-middle, lower-middle, and low-income countries) and selected countries are shown in Figure 1. We selected two countries of the largest population in each income group to represent the corresponding income group. So we got 8 countries including the United States and Japan to represent the high income group; China and Brazil to stand for the upper-middle income group; India, Indonesia and Bangladesh, Nigeria to represent the lower-middle and low income group respectively.

**Figure1.**
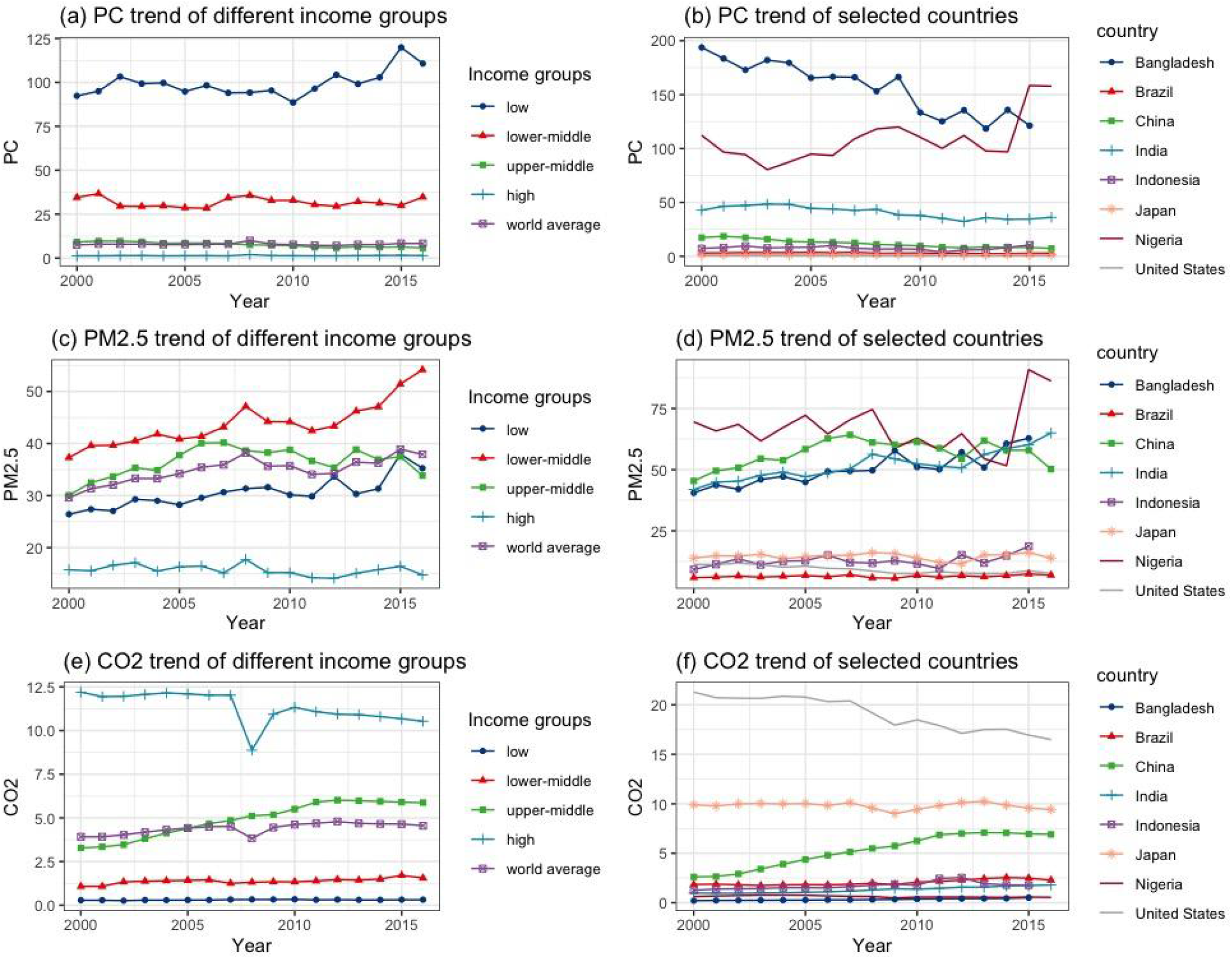
PC trends of the whole world, different income groups and selected countries. **Notes:** PC: PM_2.5_ concentration per unit per capita CO_2_ emission. We used population-weighted PC, PM_2.5_ and CO_2_ to show time tendencies of different income groups. The units of PC, PM_2.5_ and CO_2_ are µg/m^3^per tonne, µg/m^3^and tonne respectively.

Globally, the average PC remained almost stable from 2000 to 2016 worldwide. PC in low income group showed an increased tendency while the upper-middle income group’s PC decreased. World-average PM_2.5_ increased with the most increment in lower-middle groups. PM_2.5_ in high income countries remained the least and kept almost flat. As for the annual average CO_2_ emission per capita trend, the world average increased by year. The high-income group took the largest part of CO_2_ emission. However, we could see the decreasing trend of CO_2_ in the high-income group. Meanwhile the low income group emitted the least and stable CO_2_. CO_2_ emission of upper-middle and lower-middle income groups increased from 2000 to 2016, too.

From 2000 to 2016, PC in Bangladesh decreased significantly (from 193.75 µg/m^3^ per tonne to 106.08 µg/m^3^ per tonne) while Nigeria increased (from 112.24 µg/m^3^ per tonne to 157.84 µg/m^3^ per tonne). By contrast, PC kept almost stable during the study period in the United States (from 0.53µg/m^3^ per tonne to 0.46 µg/m^3^ per tonne) and Japan (from 1.40 µg/m^3^ per tonne to 1.47 µg/m^3^ per tonne). The similar increasing trend of PM_2.5_ concentration could be seen in most selected countries. While the two high income countries like the United States (11.3 µg/m^3^ in 2000 and 7.6 µg/m^3^ in 2016) and Japan (13.9 µg/m^3^ in both 2000 and 2016) showed decreasing or stable trend. The United States(21.28 and 16.48 tones per capita in 2000 and 2016) and Japan(9.90 and 9.43tones per capita in 2000 and 2016) are the largest two CO_2_ emission countries among the 8 countries while Bangladesh(from 0.21 to 0.52 tones per capita) and Nigeria(from 0.62 to 0.55 tones per capita) the least.

The spatial distributions of PC during 2000 and 2016 are presented in Figure 2. In 2000, PCs in the countries like America, Europe, Australia and most countries in South America were lower than 5 (µg/m^3^ per tonne). In developing countries like China and India, PCs were higher than 10 (µg/m^3^ per tonne) but lower than 50 (µg/m^3^ per tonne). But in poor countries in Africa, most PCs were over 100 (µg/m^3^ per tonne). Specifically, PCs in Niger, Democratic Republic of Congo were over than 1000 (µg/m^3^ per tonne) and Chad over 2000 (µg/m^3^ per tonne). In 2016, PC almost showed the same spatial distribution globally. PC in China declined to 7.26 (µg/m^3^ per tonne) in 2016. PCs in Chad and Niger declined a lot but still over 1000 (µg/m^3^ per tonne). PCs in most countries of the world decreased in the past 17 years. The most remarkable decreases were observed for countries in Africa like Chad, Democratic Republic of Congo and Niger, then China and India. Meanwhile, some African countries suffered the PC growth, such as Somalia, Eritrea and Nigeria.

**Figure 2.**
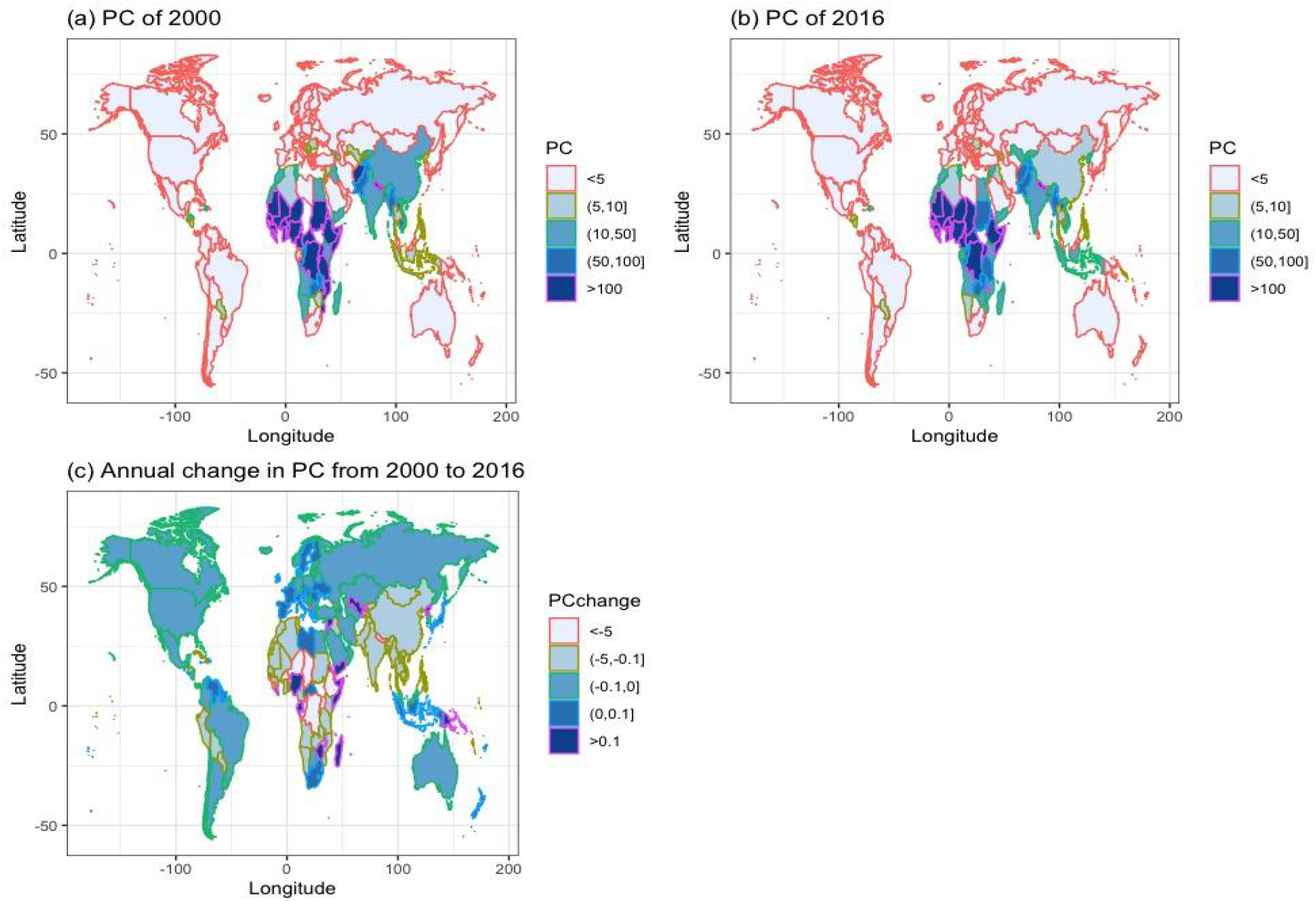
Country-level PC and annual average change in PC from 2000 to 2016. **Notes:** PC: PM_2.5_ concentration per unit per capita CO_2_ emission.The unit of PC is µg/m^3^ per tonne.

### 3.3 The relationship between PC and health metrics

The Pearson correlation coefficients between PC and LE, IMR, U5MR and LPGDP were –0.566, 0.646, 0.659, –0.585 respectively (Table S2), and all coefficients were statistically significant at the level of 0.001. Using GAMM, we investigated seven models to estimate the relation between PC and health (Table S3). In model with PC as the only independent variable, the adj.R^2^ were 0.320, 0.417 and 0.435 indicating PC independently explained 32.0%, 41.7% and 43.5% of the variation of LE, IMR and U5MR respectively. While in model with PM_2.5_ or CO_2_ as the only independent variable, PM_2.5_ and CO_2_ could only explain 3.45%, 7.81%, 10.49% and 22.11%, 22.39%, 19.84% of the respective variations of LE, IMR and U5MR. Therefore, PC seemed to be a better indicator to reflect health compared with PM_2.5_ and CO_2_. PGDP single could reflect variation of LE, IMR and U5MR by 58.0%, 63.6%, 61.3% respectively.

We examined the nonlinear associations of PC with LE, IMR, U5MR and LPGDP in Figure 3 using GAMM. We got the reverse relation curves between PC and LE, LPGDP. Simultaneously, we found a positive relation between PC and IMR, U5MR. The non-linear relationships changed minimally when we altered the covariates of the model (Figure S1).

**Figure 3.**
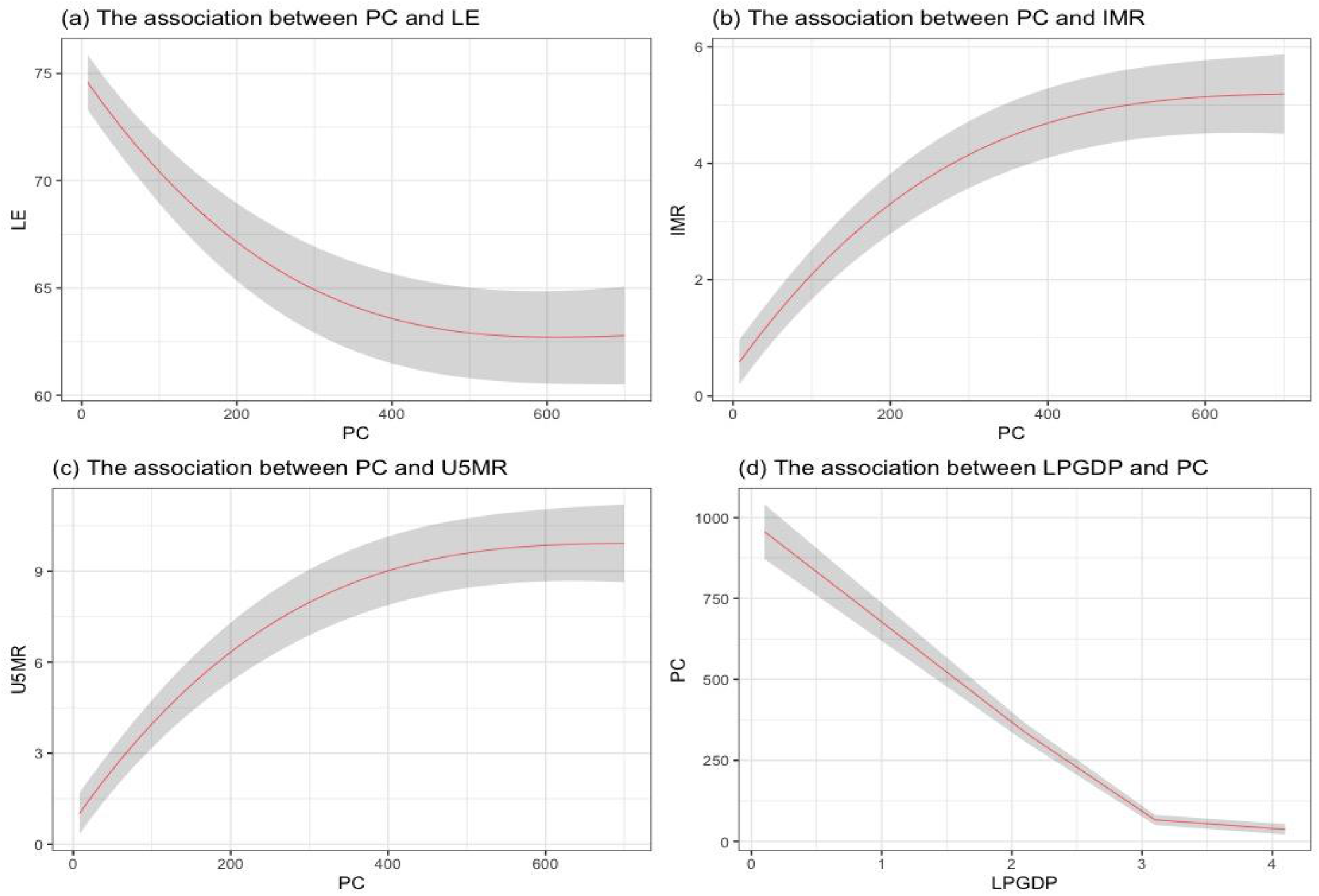
The modeled associations of PC with LE, IMR, U5MR and PGDP, by GAMM. **Notes:** Black shadow indicates 95% confidence interval (CI). LE: life expectancy at birth, IMR: Infant-mortality rate, U5MR: Under-five mortality rate, PC: PM_2.5_ concentration per unit per capita CO_2_ emission, LPGDP: logarithm of GDP per capita. The GAMMs were (a): PC+PGDP+T2M+QV2M+PM_2.5_+CO_2_; (b): PC+PGDP+T2M+QV2M+PM_2.5_+CO_2_; (c): PC+PGDP+T2M+QV2M+PM_2.5_+CO_2_; (d): LPGDP.

## 4. Discussion

To the best of our knowledge, this is the first paper to evaluate PC as a potential new indicator of air quality control efficacy. This indicator almost kept stable over 2000–2016 in the world. There is great spatial variation or inequality of PC among countries. On average, PC was high in Africa and low in America, Europe and Australia, while Asia was in the middle range during 2000–2016.

Generally, PC is decreasing as the GDP per capita grows. PC is smaller in high income or developed countries than in low income or developing countries, possibly because the use of clean-polluting production technologies increases with economic development [30]. For high income countries, they have the least PC with the highest CO_2_ emission but lowest PM_2.5_ concentration. Both PM_2.5_ concentration and CO_2_ emission showed decreasing tendency from 2000 to 2016, so there is a clear plateau for most high-income countries over the past years. Taking the United States as an example, since the 1970s the United States government has input $25 billion per year to the improvement of ambient air quality[31]. Over half of the coal-fired capacity in the United States will be equipped with the air pollution control technologies including selective catalytic reduction, electrostatic precipitators, sorbent injection and flue gas desulfurization or other scrubber technologies by 2020[32].

PC in upper-middle income countries decreased with the increase of CO_2_ and relatively slow increase of PM_2.5_. From 2000 to 2016, the decreasing PC in upper-middle groups might be contributed by technological improvement and green production promotion[30]. As the largest population country in the world and the largest upper-middle income country, PC in China decreased significantly, from 17.39 (µg/m^3^ per tonne) to 7.26 (µg/m^3^ per tonne). As the largest coal-consuming country in the world[12], the Chinese government has implemented many air quality plans such as “Air Pollution Prevention and Control Action Plan” [33] and “Reformation and Upgrading Action Plan with ultra-low emissions (ULE) technologies” focusing on controlling emissions from coal consumption, which have dramatically reduced PM_2.5_ emissions from coalfired power plants [12]. Therefore, PM_2.5_ in China remained almost unchanged from 49.5 µg/m^3^ in 2000 to 50.2 µg/m^3^ in 2016, although CO_2_ emission in China increased a lot from 2.61 tones per capita to 6.91 tones per capita.

Lower-middle income countries, most located in South Asia, PC remained almost no change from 2000 to 2016 because of both increment of PM_2.5_ and CO_2_. PM_2.5_ concentrations in South Asia mainly due to combustion emissions(solid fuels, power plants, agricultural and other open burning, industry and transportation)[34]. Taking India, the largest population country of lower-middle income and one of the highest polluted countries globally as an example [35], the major source of ambient particulate matter pollution is coal burning [36]. Although Indian government has launched several initiatives including improving technologies of coal power plants, energy-intensive industries in the past few years to reduce air pollution [37], which reduced PC in India from 42.85 (µg/m^3^ per tonne) to 36.20 (µg/m^3^ per tonne), PM_2.5_ increased from 44.9 µg/m^3^ to 65 µg/m^3^ with CO_2_ increased from 0.98 tones per capita to 1.80 tones per capita during 2000 and 2016.

Low income countries are just on the contrary to the high income ones, which had the highest PM_2.5_ concentration but lowest CO_2_ emission. PM_2.5_ increased while CO_2_ almost unchanged during 2000 to 2016, causing PC increased. The three largest PC located in the three African countries of Chad, Niger and the Democratic Republic of Congo. It is needed to mention that air pollution in Africa, such as countries in north (Niger, Egypt and Mauritania) and west (Cameroon, Nigeria and Burkina Faso) Africa and the Middle East (Saudi Arabia, Qatar and Kuwait), PM_2.5_ is typically composed of aeolian dust and vegetation fires[38,39]. Besides, 26% of 51 million people relied on biomass fuel, gas and paraffin for cooking and 41.2% for heating in the 2011 South African Census report, which will also cause the air pollution[40]. In South Africa, some policies have been promulgated such as the National Environmental Management Air Quality Act (2004) which defined the Minimum Emissions Standards for regulating gaseous and particulate emissions from industrial operations. In 2009, South Africa pledged a target of CO_2_ emissions reductions also reduced PM_2.5_ by switching away from an fossil fuels based economy[41]. PC in Chad decreased from 2286.39 µg/m^3^ per tonne in 2000 to 1163.79 µg/m^3^ per tonne in 2016 and Niger from 1496.35 µg/m^3^ per tonne to 1029.71µg/m^3^ per tonne. But the PC reduction mainly depend on the increment of PM_2.5_ (from 48.2 µg/m^3^ to 58.7 µg/m^3^ in Chad and 91.3 µg/m^3^ to 111.3 µg/m^3^ in Niger) and more fast increasing speed of CO_2_ (from 0.02 tones per capita to 0.05 tones per capita, from 0.06 tones per capita to 0.11tones per capita respectively). However, it is needed to mention that some African countries suffered the PC growth, such as Somalia, Eritrea and Nigeria. There is still a long way to go for low income countries to improve the air pollution control efficiency as part of development of economy.

PC might be a good indicator of health. PM_2.5_ attributed mortality of childhood in sub-Saharan Africa (such as Chad, Sudan, and Nigeria) and south Asia (such as India and Pakistan) contributes substantially to the global YLLs (Years of life lost) from ambient air pollution[38,39]. Meanwhile, most largest PC located in the above two areas. It was estimated that highest rate of childhood mortality due to air pollution especially PM_2.5_ was in Chad (located in sub-Saharan Africa) with the largest PC in the world (mean of PC from 2000 to 2016 was 1333.10 µg/m^3^ per tonne)[41]. In Chad, YLLs per capita due to exposure to PM_2.5_ in children younger than 5 years are 1000 times higher than in the United States(mean of PC from 2000 to 2016 was 0.48 µg/m^3^ per tonne)[39]. Meanwhile, PC might be a better indicator for monitoring national progress of addressing air pollution related health burden than PM_2.5_[2,42]or CO_2_ for the better explaining variation of LE, IMR and U5MR.

Compared with previous literature about association between PM_2.5_, CO_2_ and health[4,7], our paper suggests that more attention should also be paid to the air quality control efficacy, in order to realize climate, air quality and health co-benefits. The air pollution control efficiency could be improved through change of energy structure (e.g., shift to cleaner energy) and technology innovations (e.g., electric vehicle) [43,44]. We found that the association between PC and health metrics was independent of GDP per capita. This suggests that clean air brought by reducing PC might generate health improvements independent of economic growth. This result also suggest that the global health inequity is not merely explained by income inequality, but also by the inequality in the ability to control ambient air pollution.

Our findings contribute to the area of air pollution, climate change and human health. Firstly, it is useful for policymakers to pay more attention to air pollution control efficacy when dealing with climate change by reducing carbon emission. Secondly, PC provides a new angle to understand the global health equity. The low health levels of low income countries might be partly because of the low efficacy to reduce the harm from ambient air pollution [37]. Thus for low income countries, the promotion of air pollution control efficacy should be included as an important part of economic development. Also, assistance from developed countries to undeveloped ones should include not only improving the economy but also technologies related to air pollution control efficacy. These suggest that we could improve health equity more effectively by paying more attention to air pollution control efficiency.

The study has some limitations. Firstly, we did not obtain data from every country in the world like other global analysis[26]. Our study did not cover the Greenland, Antarctica and some Middle Africa because of the missing data. But as few people live in these areas, we could provide a reference for the majority of population in the world [26]. Secondly, due to data unavailability, we did not include data on factors that might contribute to PC such as energy structure and technologies of processing air pollution emissions. Future studies with relevant data could give a detailed evaluation on these contributing factors. There are some weaknesses of the PC index. Firstly, it couldn’t reflect the air pollution caused by the natural sources of aeolian dust and vegetation fires from the unpaved roads or deserts. Secondly, PC maybe not change while some improvements both happens in air pollution control and reducing CO_2_ per capita. That is why PC in high income countries keep stable from 2000 to 2016 as decrease happened in both PM_2.5_ concentration and CO_2_ emission. Thirdly, in theory PC would reduce if CO_2_ emission increases without impacting on ground level PM_2.5_ exposure within country. This is clearly not a good outcome to climate change and health. Anyway, PC is really a good indicator to reflect air pollution control efficiency because it reduces with changing the energy structure from coal to clean energy[33,35], improving air cleaning technology[10]. There are many ways to develop the PC indicator in the next stages. Other detailed covariates needed to be included like fossil fuel combustion emission control technology, unusual events like bushfire, natural sources and social disruptions.

## 5. Conclusions

In summary, our study developed a novel air pollution control efficacy indicator, ground level PM_2.5_ concentration per unit CO_2_ emission per capita (PC), to assess population air pollution exposure level related to carbon emission. The results indicated that PC has kept almost stable from 2000 to 2016 globally with the low income groups increased. PC is geographically different and getting lower with the economic development. PC is statistically associated with LE, IMR and U5MR, which provides a new way to promote global health equity from the angle air pollution control efficacy.

## Data Availability

The data that support the findings of this study are available upon request from the authors.

## Acknowledgements

Some data were obtained from the NASA Langley Research Center POWER Project funded through the NASA Earth Science Directorate Applied Science Program. CH was supported by Shandong provincial department of education funded projects for overseas study; SL was supported by an Early Career Fellowship of the Australian National Health and Medical Research Council (number APP1109193); and YG was supported by Career Development Fellowships of the Australian National Health and Medical Research Council (numbers APP1163693).

## Data Availability Statement

The data that support the findings of this study are available upon request from the authors.

---

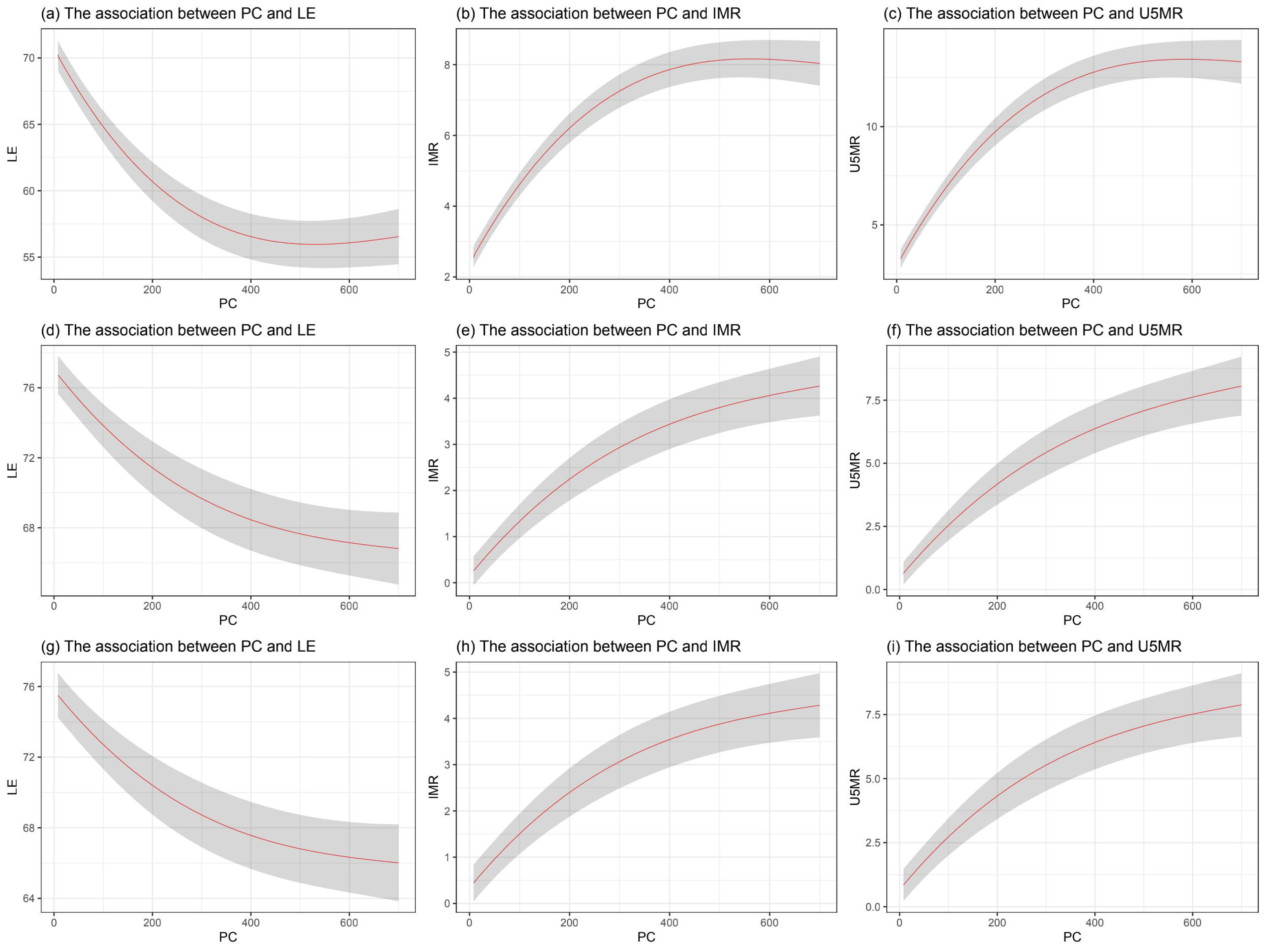

